# Glutamate Carboxypeptidase II (GCPII)-Targeted PET to Identify Muscle Denervation in Peripheral Nervous System Injuries

**DOI:** 10.64898/2026.03.18.26348533

**Authors:** William M. Padovano, Rachana Suresh, Emma K. Rowley, Aidan S. Weitzner, Maas A Khan, Keith Kuo, Zachary H. Zamore, Zohra V. Aslami, Erica B. Lee, Peter Pietri, Catherine Rutledge, Yu Su, Santosh K. Yadav, Andrew G. Horti, Ahmet Hoke, Ala Elhelali, Barbara Slusher, Catherine A. Foss, Martin G. Pomper, Sami H. Tuffaha

**Affiliations:** Department of Plastic and Reconstructive Surgery, Johns Hopkins School of Medicine; Baltimore, Maryland, United States; Department of Neurology, Johns Hopkins School of Medicine, Baltimore, Maryland, United States; Johns Hopkins Drug Discovery, Johns Hopkins School of Medicine, Baltimore, MD 21205, USA; MRB Molecular Imaging Service Center and Cancer Functional Imaging Core, Johns Hopkins University; Baltimore, Maryland, United States; Russell H. Morgan Department of Radiology and Radiological Science, Johns Hopkins University; Baltimore, Maryland, United States

**Author notes:** University of Texas Southwestern Medical Center, Dallas, Texas, United States (Martin G. Pomper). **One Sentence Summary:** Positron emission tomography (PET) targeting expression of glutamate carboxypeptidase II (GCPII) within muscle can identify denervation and reinnervation after nerve injury and repair.

## Abstract

Management of peripheral nervous system (PNS) neuropathies, such as traumatic peripheral nerve injury (PNI), relies on accurate assessment of muscle denervation and recovery. Yet, the current gold-standard clinical test, needle electromyography (EMG), has multiple shortcomings that can complicate surgical treatment. Here, we introduce a noninvasive method for holistic evaluation of muscle denervation by utilizing positron emission tomography (PET) to quantify expression of prostate-specific membrane antigen (PSMA), also known as glutamate carboxypeptidase II (GCPII), within muscles. We identified that GCPII is persistently over-expressed in denervated muscles and that expression normalizes with muscle reinnervation. Leveraging this phenomenon, we used two PSMA/GCPII-PET agents that are FDA-approved for prostate cancer imaging, [^18^F]DCFPyL and [^68^Ga]PSMA-11, to detect muscle denervation and subsequent reinnervation in experimental models of PNI. We found that denervated muscle had approximately twice the uptake as innervated muscle on GCPII-PET/magnetic resonance (MR) imaging and GCPII-PET/computed tomography (CT), which persisted for at least 16 weeks after nerve injury without repair in rats and swine. GCPII-targeted uptake also declined to near baseline levels with muscle reinnervation after nerve repair. To assess clinical feasibility, we performed [^18^F]DCFPyL PET/CT in a patient who had sustained a unilateral radial nerve injury 15 weeks prior, and we observed elevations in denervated muscle uptake that mirrored our preclinical findings. Our consistent findings across species of increased GCPII-PET uptake in chronically denervated muscle and its decline with muscle reinnervation, along with the established safety profile of available GCPII-PET agents, support the promise of GCPII-PET as a rapidly translatable strategy for characterization and longitudinal monitoring of PNIs and non-traumatic PNS neuropathies.

## INTRODUCTION

Peripheral nervous system (PNS) neuropathies, including traumatic peripheral nerve injury (PNI), often cause debilitating muscle paralysis (*1*). Understanding the extent and distribution of muscle denervation is necessary to establish the correct diagnosis, localize PNS lesions, monitor for disease progression or resolution, and determine the correct treatment strategy. Early detection of denervation is also necessary to maximize functional outcomes, as prolonged denervation leads to progressive muscle atrophy that permanently diminishes recovery potential if not addressed early (*2*).

While needle electromyography (EMG) is the current gold-standard clinical test for muscle denervation (*3, 4*), it has several important limitations. Needle EMG involves percutaneous placement of electrodes into specific muscles of interest based on surface landmarks, which can be inaccurate when testing small confluent muscle bellies in the extremities or high-risk when testing muscles near critical structures (e.g., intercostal muscles overlying parietal pleura) (*5, 6*). Assessment of volitional muscle activity depends on patient cooperation, which is often compromised in young or incapacitated patients well as those who experience anxiety and discomfort from needlesticks (*7-10*). Indeed, because needle placement is painful and testing is time-intensive, it is impractical for the tester and intolerable for the patient to evaluate all potentially involved muscles (*11*). This introduces sampling errors that can obscure diagnoses, particularly in the setting of multifocal or proximal PNS lesions (*12, 13*). Variability in testing equipment and conditions additionally complicate direct comparison between labs (*14*), and the interpretations of data obtained from EMG are largely subjective, highly operator-dependent, and cannot be independently corroborated once testing is complete (*15*). Consequently, patients referred for nerve reconstruction often require repeat testing by a specific neurologist or lab that the nerve surgeon trusts, which imposes additional costs, inconvenience and pain for patients, and potentially delays time-sensitive treatment. Given the limitations of EMG, there is a critical need to develop new diagnostic tools that provide accurate, objective, and holistic evaluation of muscle denervation to better guide the treatment of PNS pathologies.

Glutamate carboxypeptidase II (GCPII), also termed prostate-specific membrane antigen (PSMA), is an established biomarker for prostate cancer (*16*). Multiple PSMA/GCPII-targeted imaging agents are currently used in positron emission tomography (PET) for prostate cancer diagnosis, such as [^68^Ga]PSMA-11 and [^18^F]DCFPyL (*17, 18*). GCPII is expressed in several tissues outside of the prostate as well (*19, 20*), although its activity is typically very low in normal adult skeletal muscle (*21-24*). GCPII inhibition was recently shown to delay muscle loss in preclinical models for amyotrophic lateral sclerosis (ALS) and age-related atrophy, both of which involve skeletal muscle denervation with increased GCPII expression (*25-27*). We therefore hypothesized that denervation after PNI would increase muscle GCPII expression, and that this phenomenon could be noninvasively detected using GCPII-targeted imaging agents.

Here, we assessed use of GCPII-PET to provide holistic evaluation of muscle denervation. We first examined muscle GCPII expression and uptake of GCPII-targeted near-infrared (NIR) and PET agents within denervated and reinnervated muscles in rat PNI models. To assess translatability, we then performed [^68^Ga]PSMA-11 PET/CT in a porcine median nerve transection model. Finally, we performed [^18^F]DCFPyL PET/CT in a patient 15 weeks after she sustained a complete proximal radial nerve injury. Our findings demonstrate the effectiveness of GCPII-PET for holistic detection of muscle denervation and subsequent reinnervation, offering the promise of an objective and noninvasive test that addresses critical limitations of EMG for patients with traumatic PNI and other PNS neuropathies.

## RESULTS

### GCPII is expression and activity is elevated in denervated skeletal muscle

We first evaluated histologic GCPII expression in Lewis rat muscles that were subjected to either sustained denervation (i.e., nerve transection without repair) or to denervation with subsequent reinnervation (i.e., nerve transection with immediate repair). In repaired animals, neuromuscular junction (NMJ) reinnervation occurred between four and eight weeks of injury, as evidenced by co-localization of axons with motor endplates (**Fig. 1**). Small clusters of GCPII staining were noted at innervated NMJs within naïve muscle and reinnervated muscle following nerve repair but were absent at denervated NMJs following nerve injury without repair or soon after repair, before muscle reinnervation occurred. Even within innervated muscle samples, NMJs accounted for a minority of total GPCII staining. Instead, GCPII staining primarily localized to the myocyte cell membrane in both denervated and innervated myocytes, adjacent to clusters of perinuclear subsarcolemmal mitochondria (**Fig. 2**).

**Figure 1:**
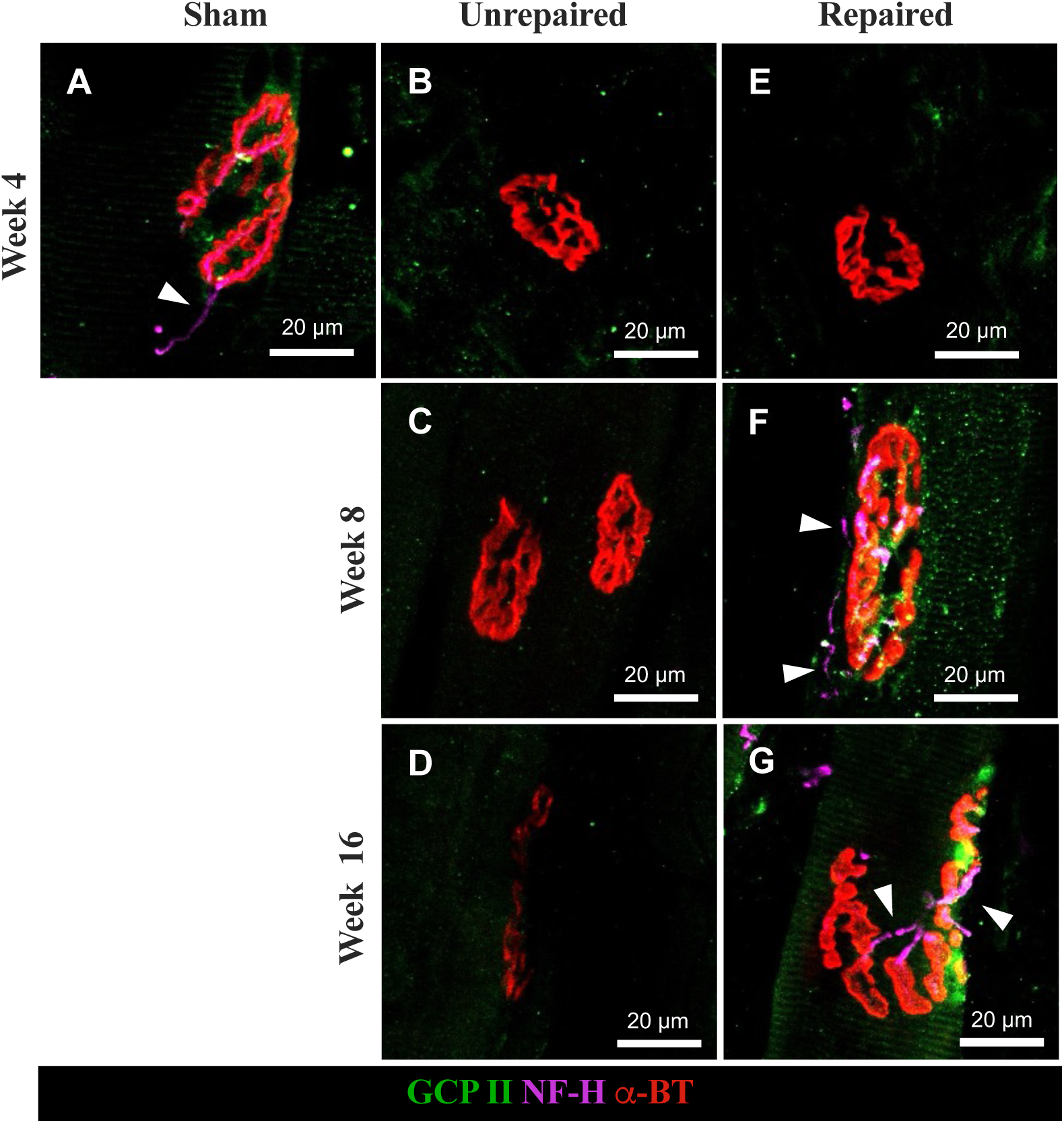
GCPII expression near innervated neuromuscular junctions. Composite images of α-bungarotoxin (red) targeting postsynaptic neuromuscular junctions (i.e., motor endplates), anti-neurofilament H (purple) targeting the axonal cytoskeletons, and anti-GCPII (green) demonstrating muscle innervation after nerve transection with or without repair; arrowheads emphasize axons. **(A)** Healthy muscle after sham surgery demonstrating a normal innervated neuromuscular junction with surrounding GCPII expression. Contrast this with the appearance of neuromuscular junctions at **(B)** 4 weeks, **(C)** 8 weeks, and **(D)** 16 weeks after sciatic nerve transection with repair; note progressive flattening and fragmentation of endplates consistent with chronic denervation, the absence of axons, and lack of GCPII expression. Nerve transection with immediate repair demonstrates infiltration of axons and neuromuscular junction reinnervation between **(E)** 4 weeks and **(F)** 8 weeks. Also note recovery of GCPII expression near the reinnervated neuromuscular junction, which persists at **(G)** 16 weeks after nerve repair. Panels are 20× magnification, scale bar: 20 μm.

**Figure 2:**
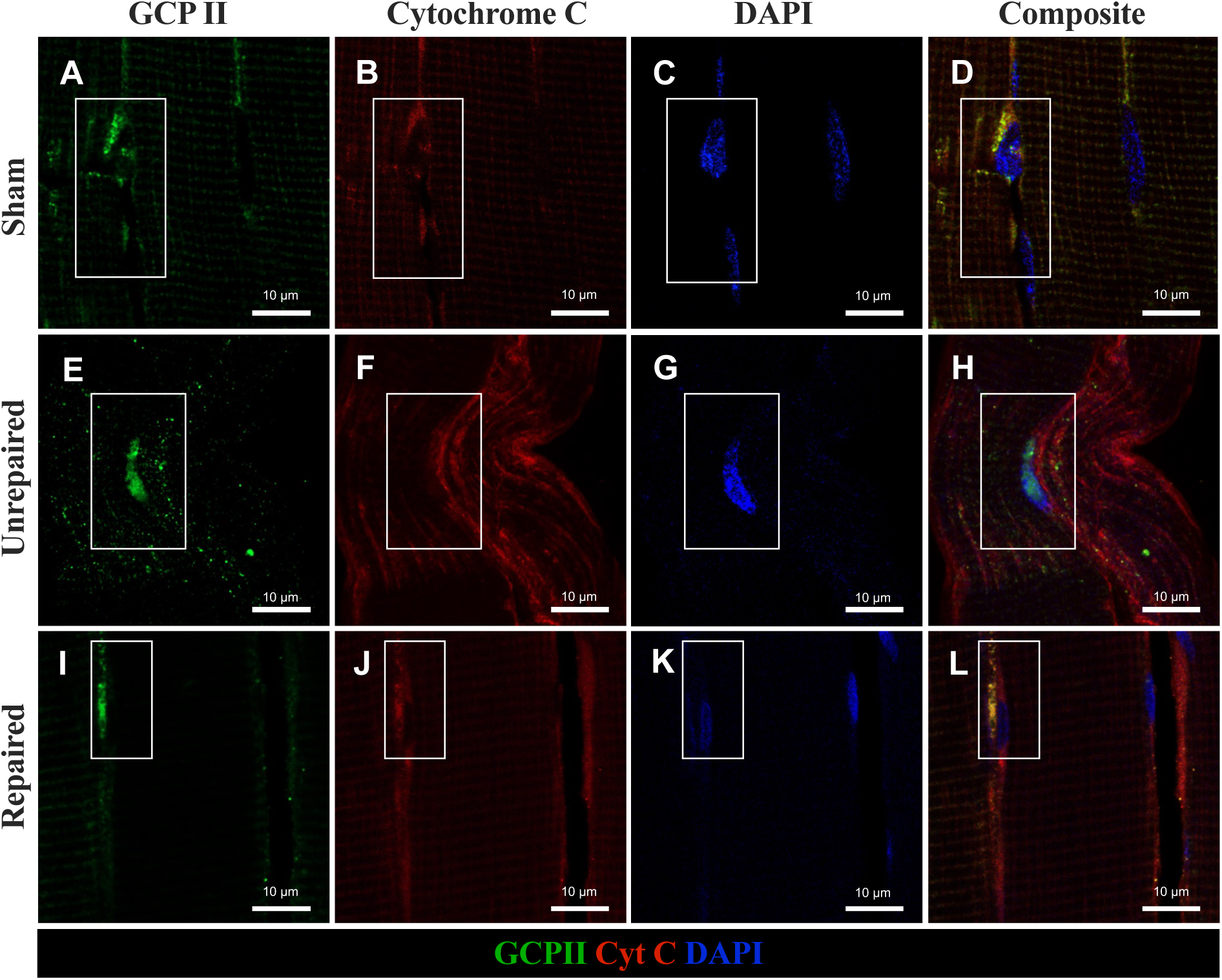
Perinuclear GCPII expression within both denervation and innervated myocytes. Images of anti-GCPII (green), anti-cytochrome C (red) targeting mitochondria, and DAPI (blue) targeting nuclei at 12 weeks after nerve transection with or without repair. White boxes around key area of each image. Panel D (sham surgery) is a composite of Panels A-C; Panel H (sciatic nerve transection without repair) is a composite of Panels E-G; Panel L (nerve transection with repair) is a composite of Panels I-K. Note localization of GCPII staining near clusters of subsarcolemmal mitochondria overlying myocyte nuclei, irrespective of denervation. A similar staining pattern was present at all tested timepoints between two and 16 weeks after nerve transection with or without repair. Panels are 63× magnification, scale bar: 10 μm.

To quantify changes in GCPII expression, we next performed Western blot after 2-, 8-, 12-, and 24-weeks of denervation. Densitometric analysis revealed persistently elevated GCPII concentration in denervated muscles relative to naïve, contralateral control muscles (**Fig. 3A**). This was corroborated by GCPII enzymatic activity assay performed on the same muscles, which demonstrated significantly elevated GCPII activity in denervated samples after 24 weeks of denervation **(Fig. 3B**). When pooled across timepoints, denervated muscles had 4.0 times higher expression on Western blot (control: 1.00 ± 0.00, denervated: 4.00 ± 2.28, mean ± SD; *P* = 0.002) and 4.3 times higher GCPII activity (control: 7.82 ± 8.40, denervated: 33.34 ± 13.81, mean ± SD; *P* <0.0001) than controls.

**Figure 3.**
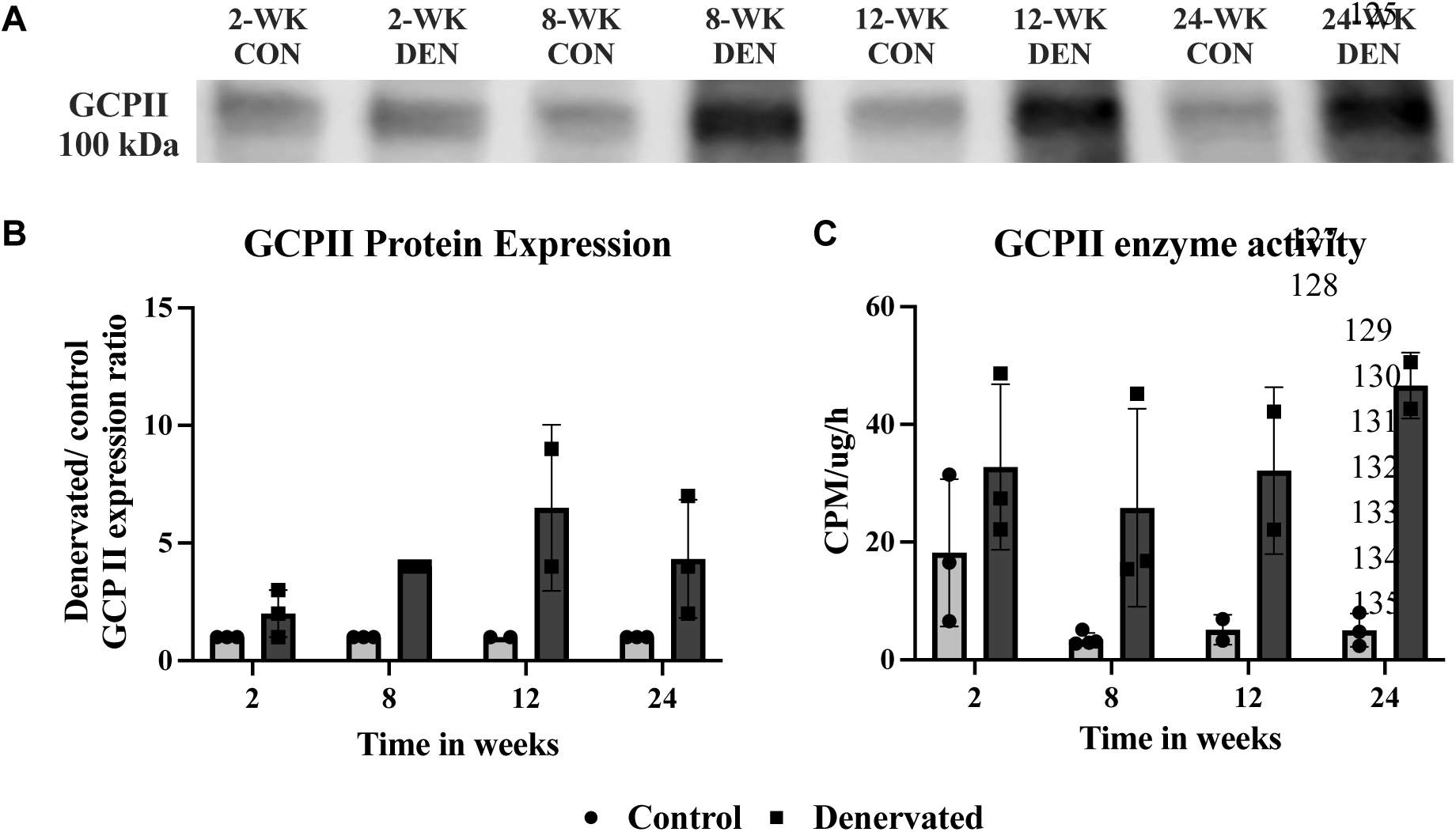
GCPII expression and activity in denervated rat lateral gastrocnemius muscle. **(A)** Representative Western blot and corresponding densitometric analysis (n=2–3 per group) show increased GCPII band intensity in denervated samples at 2-, 8-, 12-, and 24-weeks post-denervation, compared to low baseline expression in naïve muscle (i.e., control). Band intensities were normalized to denervation-duration-matched controls and plotted as relative expression. Statistical analysis was not performed due to the limited sample size. **(B)** GCPII enzymatic assay demonstrating significantly increased GCPII activity at 8-, 12-, and 24-week timepoints. Activity is expressed in femtomoles of N-acetyl-aspartyl-glutamate (NAAG) hydrolysis per mg GCPII per hour (fmol/mg/h).

### Near-infrared (NIR) imaging with YC27 distinguishes denervated and reinnervated muscles after nerve injury

We next assessed whether uptake of GCPII-targeted agents was also elevated within denervated muscles. We first evaluated YC27, a fluorescent compound that specifically binds to GCPII and emits at 800 nm, avoiding potential competing endogenous fluorescence at shorter wavelengths (*28*). We generated diverse injuries to the sciatic nerve and its distal branches in Lewis rats to examine the selectivity of YC27 uptake for denervated muscles, persistence over time in the absence of nerve repair, and responsiveness to varying degrees of reinnervation by regenerating axons. In this model (**Fig. S1**), a sciatic nerve transection with repair produces approximately four weeks of complete denervation in anterior and posterior compartment muscles until reinnervation begins (*29-36*), whereas a common peroneal nerve crush injury produces around two weeks of denervation in the anterior compartment only (*37-40*). At two weeks post-injury, YC27 uptake was increased within denervated muscles groups after nerve transection without repair, nerve transection with repair, and nerve crush (**Fig. 4A-C**). At four weeks post-injury, YC27 uptake remained elevated in denervated muscles after nerve transection irrespective of nerve repair (**Fig. 4D, E**), but uptake resolved in anterior compartment muscles after common peroneal nerve crush with muscle reinnervation (**Fig 4F**). By 16 weeks after sciatic nerve transection, YC27 uptake remained high only in the setting of nerve transection without repair (**Fig. 4G, H)**.

**Figure 4:**
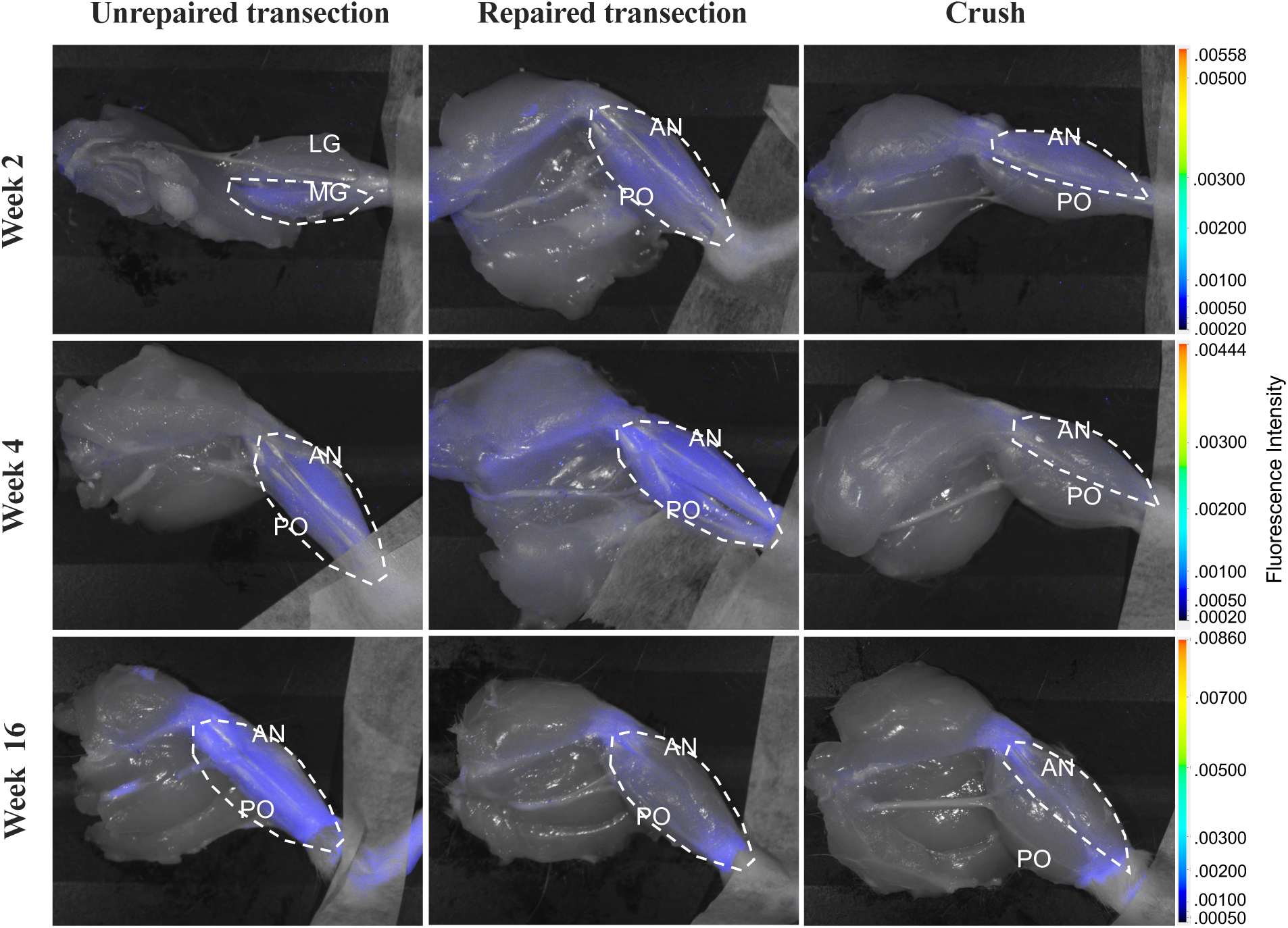
Serial near-infrared images of YC27 hindlimb uptake after various nerve injuries. Disarticulated rat hindlimbs (**A**) Transection of tibial nerve branch to the medial gastrocnemius muscle produces YC27 uptake in the medial gastrocnemius only. (**B**) Sciatic repair produces uptake in both anterior and posterior leg compartment muscles at 2 weeks. (**C**) Common peroneal nerve crush produces uptake in the anterior leg compartment muscles alone. (**D**) Sciatic nerve transection (**E**) sciatic repair both demonstrate YC27 uptake in anterior and posterior compartments at 4 weeks. (**F**) By contrast, reinnervation after common peroneal nerve crush produces reduced YC27 uptake in the anterior compartment. **(G)** By 16 weeks, sciatic transection demonstrates atrophy and YC27 uptake in chronically denervated anterior and posterior compartment muscles. **(H)** Reinnervation of anterior and posterior compartments after repair corresponds to reduced YC27 uptake. **(I)** Reinnervated of anterior compartment muscles after common peroneal nerve crush corresponds to reduced YC27 uptake. White light channel settings: minimum intensity 5.90x10^-5^, maximum intensity 5.16, gamma: 2.26. Y-axis is fluorescence intensity on 800 nm channel. Post-processing: brightness and contrast uniformly increased 30% on all images to improve visibility for reproduction (Microsoft PowerPoint V16.71). MG: medial gastrocnemius, LG: lateral gastrocnemius, AN: anterior compartment, PO: posterior compartment.

### [^18^F]DCFPyL uptake is elevated in denervated muscles

To quantify GCPII-targeted radiotracer uptake relevant for PET imaging, 48 rats (24 male, 24 female) underwent unilateral sciatic nerve transection without repair, sciatic nerve transection with repair, or sham surgery. At two, four, or 16 weeks post-injury (n=16 per timepoint), animals received 5.18 ± 1.67 MBq of [^18^F]DCFPyL intravenously prior to gastrocnemius muscle harvest. Tissue radioactivity was expressed as percent injected dose per gram of tissue (%ID/g). Gastrocnemius muscles in both repaired and unrepaired groups demonstrated increased uptake at two weeks (mean %ID/g: repaired 0.26, unrepaired 0.23, sham 0.13) and four weeks (mean %ID/g: repaired 0.31, unrepaired 0.37, sham 0.073) (**Fig. 5A**). At 16 weeks, higher [^18^F]DCFPyL uptake persisted in unrepaired animals (mean %ID/g: 0.36), but uptake declined toward sham in repaired animals (mean %ID/g: 0.19). When pooled across timepoints, mean gastrocnemius muscle %ID/g in unrepaired groups was 0.32 ± 0.13 compared to 0.12 ± 0.06 for sham (*P* < 0.0001).

**Figure 5:**
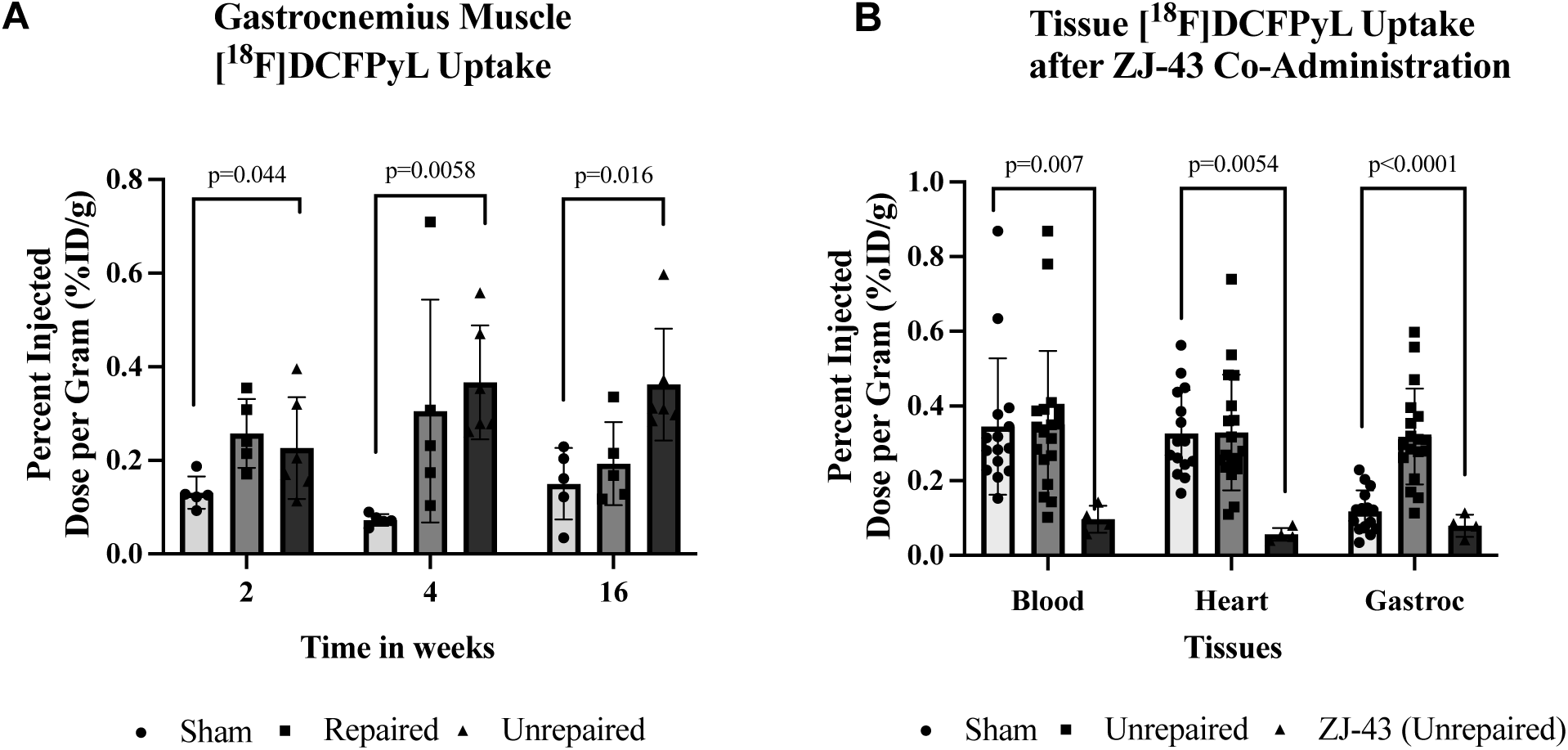
Rat [^18^F]DCFPyL biodistribution: **(A)** Mean gastrocnemius muscle uptake administration 2, 4, and 16 weeks after injury. White: sham surgery (uninjured nerve); Gray: repaired nerve; Black: unrepaired nerve; n = 5-6 per group per timepoint. Note persistent muscle uptake in the unrepaired group over time **(B)** Tissue [^18^F]DCFPyL uptake after blocking GCPII binding with ZJ-43 coadministration; White: sham surgery (uninjured nerve), pooled across all timepoints; Gray: unrepaired nerve, pooled across all timepoints; Black: ZJ-43 co-administration, 4 weeks after sciatic nerve transection without repair. One outlier in biceps muscle unrepaired group (1.2 % ID/g) was excluded in the plot for improved visibility of remaining data; the outlier was included in statistical comparisons. Groupwise comparisons performed using Kruskal-Wallis test. %ID/g: percent injected dose per gram.

### Muscle [^18^F]DCFPyL uptake is attributable to GCPII expression

To confirm that [^18^F]DCFPyL uptake in denervated muscles is specific for GCPII expression, four additional animals underwent co-injection of 3.7 ± 0.074 MBq of [^18^F]DCFPyL and 100 mg/kg of ZJ-43, a potent competitive inhibitor of GCPII binding (*41*), four weeks after sciatic nerve transection without repair. ZJ-43 administration eliminated differences in [^18^F]DCFPyL uptake between denervated and innervated gastrocnemius muscles (**Fig. 5B**).

### Sustained [^68^Ga]PSMA-11 uptake within denervated muscles on serial PET/MR in rats

To assess whether changes in muscle GCPII uptake are detectable on PET, we next performed serial [^68^Ga]PSMA-11 PET/MRI in six male Lewis rats after right sciatic nerve transection with or without repair (n = 3 per group). Animals were imaged at four weeks and 16 weeks post-injury. Consistent with [^18^F]DCFPyL biodistribution results, both groups exhibited increased uptake at four weeks (**Fig. 6**). The ratio of target (operative side) to non-target (non-operative side) SUVmean (T/NT) in affected versus unaffected hindlimbs was 2.12 ± 0.14 and 2.29 ± 0.15 for repaired and unrepaired groups, respectively. At 16 weeks, nerve repair resulted in a statistically significant decline in uptake relative to unrepaired animals (unrepaired: 1.85 ± 0.18, repaired: 1.37 ± 0.17, *P* = 0.037).

**Figure 6:**
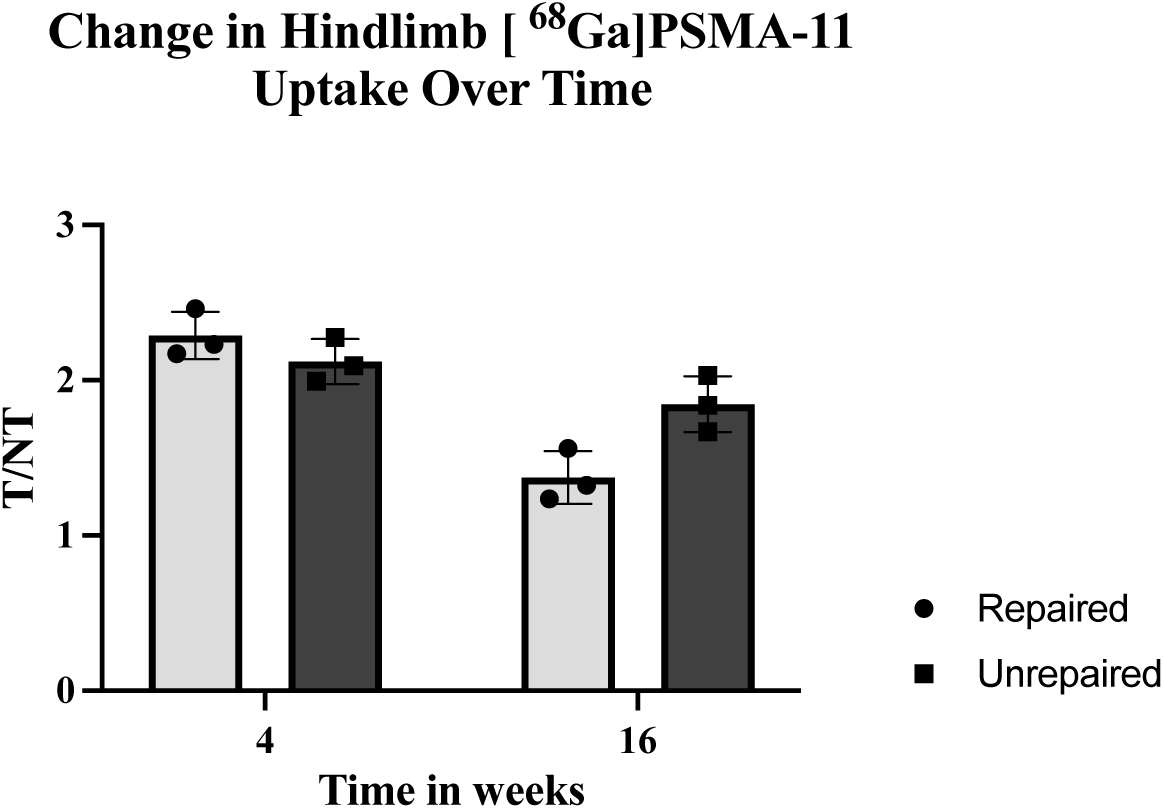
Serial rat [^68^Ga]PSMA-11 PET/MRI. The same six animals were imaged at week 4 and week 16 after right sciatic transection with or without repair. T/NT: ratio of target (right) hindlimb and non-target (left) hindlimb muscle mean activity in SUVmean. Repeated measures ANOVA demonstrated a significant change in trend between unrepaired and repaired groups (*P* = 0.037). Y-axis reference set to 1.0.

### [^68^Ga]PSMA-11 PET/CT identifies denervated muscles after median nerve injury in a porcine model

To further validate our rodent PET/MR findings in a large animal model, a female Yorkshire pig underwent bilateral median nerve transection at the proximal humerus. The median nerve diameter, regenerative capacity, and regenerative distance between injury and forearm flexor muscles are comparable to humans (*42, 43*). At 16 weeks post-injury, the animal received 89.17 MBq of [^68^Ga]PSMA-11 prior to whole-body PET/CT with a clinical scanner (**Fig. 7**). A 16-week timepoint was selected to allow for direct comparison against rat PET/MR results. The median nerve-innervated muscles were confirmed to be denervated at the time of imaging, with denervated NMJs observed on muscle histology (**Fig. S2**). Mean activity on PET/CT was elevated in all median nerve-innervated muscle groups relative to naïve muscle references (**Table S1**). When pooled across muscle groups, denervated muscles had 1.88 times greater activity than naïve muscles (SUVmean 0.49 versus 0.26).

**Figure 7.**
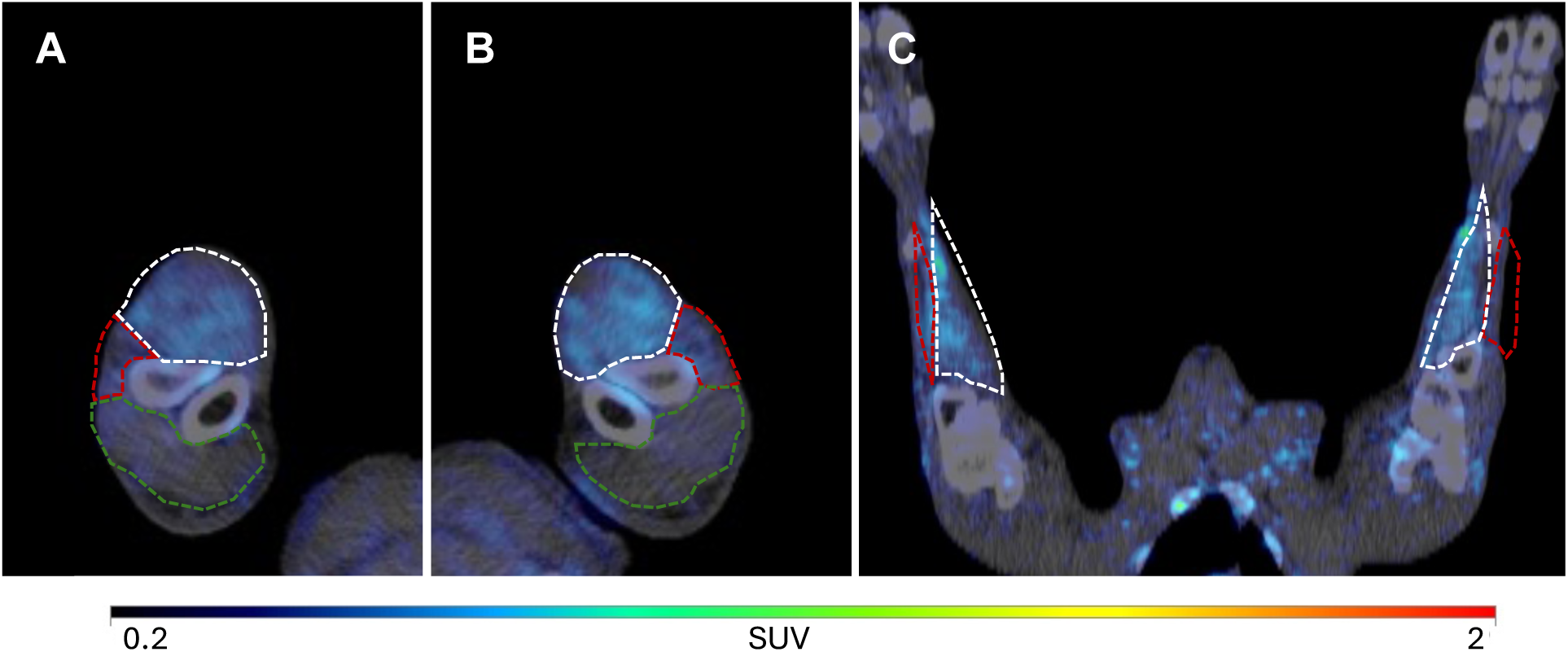
Porcine PET/CT 16 weeks after bilateral median nerve transection. Forearm muscles innervated by the median nerve circled in white, ulnar nerve circled in red, radial nerve circled in green. Axial views of **(A)** right and **(B)** left forearm demonstrating selective uptake within the denervated median nerve-innervated forearm muscles (white), consistent with selective median nerve injuries. **(C)** Coronal view of the same animal redemonstrating selective uptake within median nerve-innervated forearm muscles. Uptake expressed in SUV. The animal received 89.17 MBq of [^68^Ga]PSMA-11 one hour prior to imaging.

### Human [^18^F]DCFPyL PET/CT identifies denervated muscles after radial nerve injury

Following validation in small and large animals, we performed [^18^F]DCFPyL PET/CT in a woman who had sustained a complete left radial nerve palsy 15 weeks prior to imaging. This patient was selected for having a discrete, clinically confirmed nerve injury with similar chronicity as our preclinical injury models. Physical examination and electrodiagnostic evaluation prior to imaging confirmed absent radial nerve function and complete muscle denervation distal to the triceps muscle, including the supinator and brachioradialis muscles. She received 344.1 MBq of [^18^F]DCFPyL one hour prior to PET/CT from vertex of the head to mid-thigh (**Fig. 8)**. Consistent with our rat and pig PET results, activity in the affected radial nerve-innervated muscles was 2.0 times greater than in the contralateral arm (**Table S2)**.

**Figure 8:**
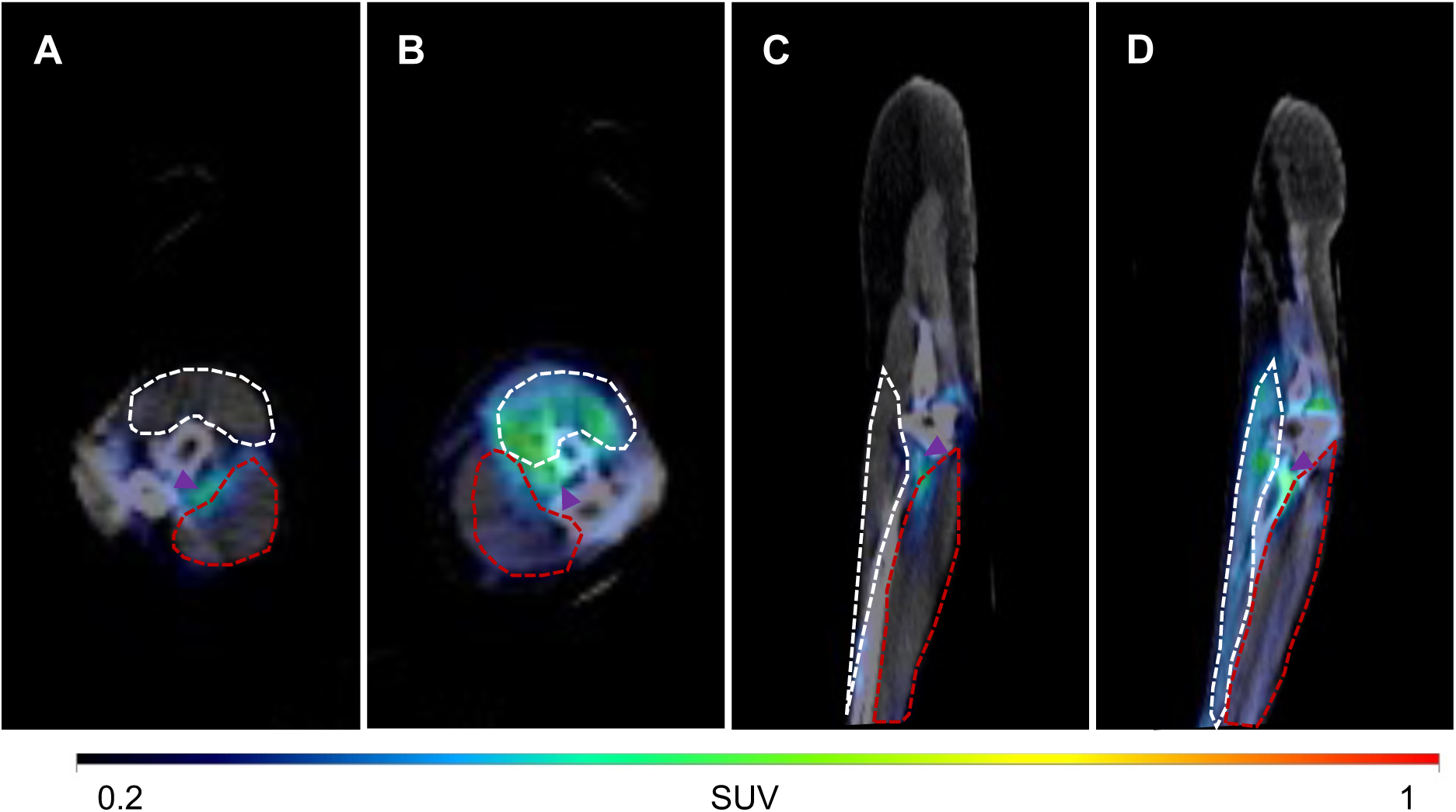
Human PET/CT 15 weeks after left radial nerve injury. The patient had no clinical or electrodiagnostic evidence of radial nerve recovery at the time of imaging. Forearm extensor compartment and mobile wad muscles, which are innervated by the radial nerve distal to the site of injury, are circled in white and forearm flexor compartment muscles, which are innervated by the median and ulnar nerves, are circled in red. Axial views of the proximal **(A)** uninjured right forearm and **(B)** injured left forearm, which demonstrate selective uptake within the left radial nerve-innervated forearm muscles (white). Sagittal views of the **(C)** uninjured right forearm and **(D)** injured left forearm, which demonstrate elevated uptake in the left radial nerve-innervated forearm extensor compartment relative to the left flexor compartment (red) and to the extensor and flexor compartments on the right side. Note focal uptake (arrowheads) near the brachial artery at the elbow, contrasted with the uniform uptake throughout involved muscles. Uptake expressed in SUV.

## DISCUSSION

The key findings of this study are that muscle denervation leads to sustained increases in muscle GCPII expression, which are detectable with clinically available PSMA/GCPII-PET agents. Muscle denervation led to increased uptake of the GCPII-targeted NIR agent YC27 and the GCPII-PET agents [^18^F]DCFPyL and [^68^Ga]PSMA-11 at all tested timepoints between two and 16 weeks post-injury. Muscle reinnervation after nerve repair also led to a reduction in uptake of YC27, [^18^F]DCFPyL, [^68^Ga]PSMA-11 toward baseline, enabling the use of GCPII-PET to monitor recovery from spontaneous regeneration or after nerve repair. Moreover, we demonstrated that the same [^18^F]DCFPyL PET protocol (i.e., dose, time interval between injection and imaging, and acquisition time) commonly used for prostate cancer imaging revealed the expected pattern of muscle denervation in a patient with a who had sustained a clinically-diagnosed radial nerve injury.

GCPII-PET addresses multiple limitations of EMG by providing quantitative, reproducible, and noninvasive evaluation of all potentially affected muscles, generating imaging data that can be independently interpreted and verified by treating physicians (*11, 14*). Our findings also suggest the superiority of GCPII-PET to fluorodeoxyglucose (FDG)-PET for diagnosis of PNIs (*68, 69*), as changes in glucose metabolism after nerve injury are heterogeneous and short-lived (*70-73*). By contrast, GCPII-PET demonstrated consistent differences between denervated and non-denervated muscles that were sustained over several months following PNI. GCPII-PET has special relevance to conditions with complex and unpredictable patterns of muscle denervation, such as proximal PNIs involving the brachial or lumbosacral plexus, Parsonage Turner syndrome (i.e., neuralgic amyotrophy), and spinal cord injury (SCI) with lower motor neuron involvement. In such cases, comprehensive evaluation of all potentially involved muscle groups will avoid the risk of sampling errors inherent in EMG (*44*).

While prior studies observed acute elevation in muscle GCPII expression in an ALS mouse model, we demonstrated that this phenomenon also occurs in denervated muscle following traumatic nerve injury, is sustained for at least four months of chronic denervation, and declines with muscle reinnervation. These characteristics make muscle GCPII expression a fortuitous biomarker to enable both diagnosis and monitoring of disease progression or recovery. The biologic rationale for increased GCPII expression in denervated muscles is not well understood. GCPII is a transmembrane glycoprotein that converts N-acetyl-aspartyl-glutamate (NAAG) into glutamate and N-acetyl-aspartate (NAA) and also liberates glutamate from pteroylpoly-gamma-glutamate to form pteroylglutamate (*53*). Glutamate signaling modulates competitive synaptic elimination at mammalian NMJs during postnatal development, and GCPII is expressed at NMJs during this period (*54-56*). Polyneuronal innervation and synaptic elimination has also been demonstrated at nascent NMJs in reinnervating adult rat muscle (*57*). However, we demonstrated that synaptic GCPII expression was only present in innervated NMJs, including in naïve muscles and reinnervated muscle following nerve repair, and thus does not explain the uniformly increased GCPII-targeted uptake in denervated muscles. Similar findings were reported by Slusher et al. in a rodent model for amyotrophic lateral sclerosis (ALS), in which the vast majority of muscle GCPII staining occurred outside of NMJs (*25*). Alternatively, increased GCPII expression may reflect a compensatory change resulting from broader alterations in protein degradation and synthesis (*58*). Denervation increases glutamate and reduces glutamine concentrations within myocytes (*59, 60*), and these changes have been linked to constitutional activation of both the ubiquitin-proteasome pathway for protein degradation and rapamycin complex 1 (mTORC1) for autophagy suppression (*61-63*). A potential link between GCPII and sustained changes in protein homeostasis after denervation is consistent with the chronic increases in both GCPII expression and GCPII-targeted agent uptake that we observed in denervated muscles. Future mechanistic investigations to elucidate the pathophysiologic underpinnings and implications of GCPII activity in denervated muscle will shed light on potential therapeutic targets to mitigate the deleterious effects of denervation-induced muscle atrophy.

This study has several limitations. For one, the persistence of GCPII-PET uptake in the setting of very long-term denervation remains unclear. Although the histologic and functional consequences of chronic denervation-induced muscle atrophy in rodents are analogous to those in humans, the rate of progression is accelerated in rats, with end-stage muscle atrophy reached with approximately seven months of denervation compared to 18-24 months in humans (*64, 65*). We evaluated muscles up to 16-24 weeks post-injury as it approximates the end of the clinically-relevant window in patients, beyond which functional muscle reinnervation becomes unlikely (*66, 67*). While our clinical PET data provides a promising proof-of-concept, additional clinical testing is needed to define the clinical sensitivity and specificity of this diagnostic modality, particularly in cases of partial denervation.

In conclusion, GCPII-PET is a promising method for non-invasive, quantifiable, and holistic characterization of muscle denervation and subsequent reinnervation, thereby addressing several unmet needs in the evaluation of PNS injury. Given the established safety and clinical availability of [^18^F]DCFPyL and [^68^Ga]PSMA-11, GCPII-PET is well-positioned for translation to patient care.

## MATERIALS AND METHODS

### Study Design

The objective of this study was to address the unmet need for noninvasive diagnosis and monitoring of complex PNS neuropathies. To meet this objective, we evaluated whether muscle uptake of GCPII-targeted PET agents could serve as a noninvasive biomarker for muscle denervation. While several proteins are elevated in denervated muscles and traumatized nerves, GCPII is a highly attractive target because GCPII-based PET is already employed clinically for prostate cancer imaging. We hypothesized that the PET agents, [^18^F]DCFPyL and [^68^Ga]PSMA-11, could be repurposed to characterize muscle innervation. We first evaluated the concentration and activity of GCPII in denervated and innervated muscles. We next assessed uptake of known GCPII-targeted agents in small- and large-animal PNI models after various nerve injuries and at multiple timepoints post-injury. We performed ex vivo NIR imaging in nerve-injured Lewis rats after intravenous YC27 injection and corroborated NIR results using a separate cohort of rats, which underwent unilateral sciatic transection without repair, transection with repair, or sham surgery. At two, four, or 16 weeks post-injury, these animals received intravenous [^18^F]DCFPyL injection prior to tissue harvest and quantification using an automated gamma counter. A third cohort of rats underwent serial in vivo [^68^Ga]PSMA-11 PET/MRI at four and 16 weeks after sciatic nerve transection with or without repair. We next examined the translatability of these results to large animals by performing [^68^Ga]PSMA-11 PET/CT in a Yorkshire pig 16 weeks after bilateral median nerve injuries. Finally, we evaluated muscle [^18^F]DCFPyL PET/CT uptake in a woman who had sustained a left radial nerve injury 15 weeks prior. For these experiments, histologic assessments were blinded to injury type. Uptake measurements of GCPII-targeted agents were unblinded, as the injuries produced overt anatomic changes to affected limbs. Animal protocols were approved by our Institutional Animal Care and Use Committee (Protocols RA21M203, RA22M257, SW23M91) and were conducted per National Institutes of Health (NIH) guidelines for animal welfare. Clinical imaging was performed after obtaining informed consent and with Institutional Review Board approval (IRB00422704).

### Synthesis of GCPII-Targeted Imaging Agents

The properties and synthesis of YC27 were described previously (*28*). In brief, YC27 is a high-affinity GCPII-binding urea (*K*_i_ 0.37 nM) that is functionalized with the commercially-available NIR dye, IRDye800CW (LI-COR Biosciences, Lincoln, NE, USA). YC27 exhibits a maximum absorbance of 774 nm and fluoresces with a maximum emission at 792 nm. For animal studies, [^18^F]DCFPyL was synthesized at the Johns Hopkins PET Center as described previously (*74*). [^68^Ga]PSMA-11 (Illuxix®; Telix Pharmaceuticals, Melbourne, Australia) was purchased from a local radiopharmacy and supplied in a 10 mL solution suitable for injection. For both radiopharmaceuticals, 100-300 µL doses of the parent solution were drawn to reach the target activity necessary for rodent and porcine evaluations. For the clinical scan, [^18^F]DCFPyL was purchased from the clinical manufacturer (Pylarify®; Lantheus, North Billerica, MA, USA).

### Animal Models

Rodent surgical procedures involved Lewis rats (Charles River, MD, USA) aged 14-16 weeks and were performed under general anesthesia via inhalational isoflurane. The rat sciatic nerve branches into a common peroneal nerve branch to anterior leg compartment muscles and a tibial nerve branch to posterior leg compartment muscles, including separate branches to the medial and lateral heads of the gastrocnemius muscle. For animals undergoing nerve transection with repair, the sciatic nerve was sharply divided using straight microsurgical scissors and immediately repaired using three 8-0 non-absorbable monofilament epineurial sutures. For sciatic nerve transection without repair, the nerve was sharply transected, and the distal stump was mobilized upward 120 degrees clockwise and tacked to nearby muscle fascia using one 8-0 non-absorbable monofilament suture to prevent spontaneous reinnervation. For common peroneal nerve crush, the common peroneal nerve was exposed at its origin from the sciatic nerve and a Webster 5″ smooth needle holder (Aesculap, MO, USA) was closed over the nerve for 30 seconds. For isolated medial head of gastrocnemius muscle denervation, the tibial nerve was identified at its origin at the sciatic nerve and exposed distally between the medial and lateral heads of the gastrocnemius muscles. The branch to the medial head was isolated and segmentally resected from its origin at the tibial nerve to its muscular insertion to prevent spontaneous reinnervation. For sham surgery, the sciatic nerve was exposed and gently mobilized from the underlying muscle fascia. In all rodents, the surgical wound was closed in layers using 4-0 absorbable sutures.

The pig surgery involved a female Yorkshire pig (Archer Farms, MD, USA) aged 22 weeks. In a dedicated large animal operating room, the animal was placed supine with bilateral upper extremities abducted 90 degrees. General anesthesia was provided via endotracheal isoflurane. For each upper extremity, a longitudinal incision was made in the axilla just posterior to the anterior axillary fold. The pectorals major muscle belly was bluntly split longitudinally to expose the proximal median nerve, which was sharply transected using an 11-blade scalpel at a position 10 cm from the sternum and 12 cm from the olecranon. The pectoralis muscle was repaired, and the skin closed in layers using 4-0 absorbable sutures.

### Biodistribution

Forty-eight rats (24 male, 24 female) received right sciatic nerve transection without repair (n=18), transection with repair (n=15), or sham surgery (n=15), and underwent biodistribution studies at two, four, or 16 weeks post-injury (n=16 per timepoint). Animals received 5.18 ± 1.67 MBq of [^18^F]DCFPyL via tail vein injection two hours prior to euthanasia by isoflurane overdose and cervical dislocation. Tissues were rapidly harvested and weighed prior to quantification using an LKB CompuGamma CS 1282 automated gamma counter (PerkinElmer, Waltham, MA, USA). Tissues included the right gastrocnemius muscle (operated side), left gastrocnemius muscle (non-operated side), blood, heart, and kidney. Counts were background- and decay-corrected and compared against a 1:10 diluted standard dose to calculate percent injected dose per gram (%ID/g) for each tissue. This calculation was performed as follows, where CPM represents counts per minute:

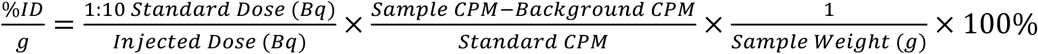

### Blocking Studies

An additional four Lewis rats underwent sciatic nerve transection without repair. At four weeks post-injury, animals received co-injection of 3.7 ± 0.74 MBq of [^18^F]DCFPyL and 100 mg/kg of the competitive GCPII inhibitor ZJ-43 (Tocris Biosciences, Bristol, UK). Biodistribution of [^18^F]DCFPyL was assessed after two hours of uptake time as described above.

### Near-Infrared (NIR) Imaging

Six Lewis rats received multiple nerve injuries, including sciatic nerve transection with or without repair, common peroneal nerve crush, and selective transection of the medial gastrocnemius muscle branch of the tibial nerve. Imaging was performed two, four, or 16 weeks post-injury. Animals received a 10 nM dose of YC27 via tail vein injection 24 hours prior to euthanasia, hindlimb disarticulation, and imaging using a Pearl Impulse Near-Infrared Imager (LI-COR Biosciences, Lincoln, NE, USA). Images were acquired with a 790/800 nm band-pass filter and overlaid on a white light photograph using the manufacturer’s software (Pearl Impulse version 2.0).

### Small Animal PET/MRI

Six rats underwent sciatic nerve transection without repair (n = 3) or transection with repair (n = 3). Animals were imaged serially using a 7T simultaneous PET/MRI scanner (Bruker BioSpec 70/30; Bruker Corporation, Billerica, MA, USA) at four and 16 weeks post-injury. An average 3.8 ± 0.51 MBq of [^68^Ga]PSMA-11 was administered via tail vein injection one hour prior to each imaging session. Scans were performed from hip to ankle using the following parameters: multislice axial TurboRARE T2-weighted MRI scan with echo time msec/repetition time msec (TE/TR) of 30/4000, resolution of 0.25 × 0.25 mm, field of view of 70 × 50 mm, 40 sections, 1-mm section thickness, matrix size of 280 × 200, and four averages. Simultaneous 10-minute static PET scans were acquired with a three-ring Bruker Si 198 PET and MRI coil using a 72 mm PET-optimized Tx/Rx radiofrequency coil (Bruker Corporation, Billerica, MA, USA) centered inside the PET detector. The acquired PET images were reconstructed using a three-dimensional maximum-likelihood expectation-maximization (MLEM) iterative image reconstruction algorithm with a pixel size of 0.5 mm and 18 iterations. The PET/MR images were spatially co-registered using a geometry-based approach with MR images as a reference in the Bruker ParaVision 360 software and converted to Digital Imaging and Communications in Medicine format (DICOM). The PET data visualizations and quantitative analysis were performed using PMOD software version 4.004 (PMOD Technologies, Zurich, Switzerland).

### Large Animal PET/CT

At 16 weeks post-injury, the Yorkshire pig underwent intravenous injection of 89.17 MBq of [^68^Ga]PSMA-11 and was imaged on a Siemens Biograph mCT PET/CT scanner (Siemens Healthineers, Knoxville, TN, USA) after one hour of uptake. PET data was collected for 70 minutes using a multi-bed dynamic protocol. The animal was positioned supine and maintained under general anesthesia via endotracheal isoflurane for the duration of the scan. Following imaging, the animal was euthanized via barbiturate overdose. PET images were reconstructed with an ordered subset expectation maximization (OSEM) algorithm, with corrections for attenuation, scatter, random, and time-of-flight (*75*). A total of two iterations with 21 subsets per iteration were used. A post-reconstruction Gaussian filtering with a filter size of 5 mm was applied to reduce noise. The software package PMOD version 3.7 (PMOD Technologies, Zurich, Switzerland) was used for PET image processing and analysis. The volumes of interest (VOIs) were manually drawn on co-registered PET/CT images. Mean standardized uptake value (SUVmean) in each VOI was calculated from the mean activity (MBq/cc) divided by the ratio of injected radioactive dose (MBq) and body mass (g).

### Human PET/CT

A woman with a left radial nerve injury at the mid-humerus resulting in absent radial nerve function distal to the triceps muscle underwent imaging 15 weeks post-injury. The patient received 344.1 MBq of [^18^F]DCFPyL and underwent Siemens Biograph Vision PET/CT (Siemens Healthineers, Knoxville, TN, USA) after one hour of uptake. The scan was performed from the vertex of the head to mid-thigh using seven bed positions, with two minutes of acquisition per bed position. This protocol is the same as is typically used for prostate cancer imaging at our center. PET images were reconstructed with an OSEM algorithm, with corrections for attenuation, scatter, random, and time-of-flight. A total of eight iterations with five subsets per iteration were used. A post-reconstruction Gaussian filtering with a filter size of 6 mm was applied. The VOIs were manually drawn on co-registered PET/CT images and SUVmean was quantified using PMOD software as described above.

### Histologic Evaluation

Immunofluorescence was performed on tissue sections fixed with 4% paraformaldehyde (PFA) and cryoprotected in 15% and 30% sucrose. Tissues were embedded in O.C.T. tissue freezing medium (ThermoFisher 23-730-571) and sectioned at 20 μm thickness (HM525 NX, Epredia). Slides were permeabilized with 0.2% Triton X-100 for ten minutes. Sections were blocked for an hour with 5% normal goat serum and 2% bovine serum albumin. Sections were incubated overnight at 4℃ with the primary antibodies of interest (see below). After washing three times with PBS, tissue sections were treated on the following day with the secondary antibodies (see below). Sections were mounted using Fluoromount G mounting medium with DAPI for nuclear staining. Confocal imaging was performed using a Zeiss LSM 800 laser scanning confocal microscope (Carl Zeiss AG, Oberkochen, Germany) equipped with Plan-Apochromat 20×/0.8 NA and 63×/1.4 NA oil DIC objectives and 405, 488, 561, and 640 nm laser lines. Images were acquired with a pinhole size set to one airy unit, bidirectional scanning, scan speed of 8, and 2× line averaging. For z-stack acquisition, the optimal step size was calculated according to the Nyquist theorem. Maximum intensity projections were generated for three-dimensional visualization. Image acquisition and processing were controlled using ZEN Blue software. Images were processed in ImageJ (NIH, Bethesda, MD, USA). The Despeckle function was applied to reduce background noise, and contrast enhancement was performed to increase pixel intensity differences between signal and background. For GCPII content, primary antibody was rabbit anti-GCPII polyclonal antibody (Proteintech, 13163-1-AP, 1:50), secondary antibody was anti-rabbit CoraLite488-conjugated IgG (H+L) (Proteintech, SA00013-2, 1:200). For mitochondrial staining, primary antibody was mouse anti-cytochrome C monoclonal antibody (Invitrogen, 45-6100, 1:100), secondary antibody was goat anti-mouse Alexa Fluor 594 IgG1 (Invitrogen, A-21125, 1:200). For motor endplates, we used alpha-bungarotoxin conjugate (Invitrogen, B13423, 1:1000). For neurite infiltration, primary antibodies were chicken anti-neurofilament H (200 kDa) antibody (Millipore Sigma, AB5539, 1:800) or rabbit β-Tubulin (Sigma Aldrich T8578, 1:1000), and secondary antibodies were goat anti-chicken Alexa Fluor 647 IgY (H+L) (Invitrogen, A-21449, 1:800) or goat anti-rabbit CoraLite488-conjugated IgG (H+L) (Proteintech, SA00013-2, 1:200).

### Western Blot

Tissue samples were dissected on ice and homogenized for 40 seconds in 150 µL of ice-cold T-PER lysis buffer (Thermo Fisher Scientific, 78510) with protease inhibitor (1:100; Thermo Fisher Scientific, 87785). Lysates were then centrifuged at 14,000 rpm for 14 minutes at 4 °C, after which the supernatant was carefully collected. Protein concentration was determined using a bicinchoninic acid (BCA) assay (Thermo Fisher Scientific, 23227), and lysates were adjusted to equal concentrations (2.5 mg/mL). Equal amounts of protein (50 µg per lane) were then combined with 6× Laemmli SDS sample buffer, heated at 70 °C for 15 min to denature proteins, briefly centrifuged, and allowed to cool at room temperature. Samples were then loaded at equal volumes (20 µL) onto 4–15% TGX precast gels (Bio-Rad, cat. no. 4561081). Electrophoresis was performed using Tris–glycine–SDS running buffer at 30 V for 30 minutes, followed by separation at 120 V for 60 minutes. Proteins were transferred onto PVDF membranes using a Trans-Blot Turbo system (Bio-Rad, 1704156EDU) at 1 A and 25 V for 30 minutes. Membranes were blocked for 5 minutes at room temperature in EveryBlot blocking buffer (Bio-Rad, cat. no. 12010020) and then incubated overnight at 4 °C with primary antibodies diluted in blocking buffer (mouse anti-GCPII, 1:500, Novus Biologicals, cat. no. NBP1-45057; rabbit anti-GAPDH (14C10), 1:30,000, Cell Signaling Technology, cat. no. 2118S). Membranes were washed with TBS-T (four quick washes followed by five, five-minute washes) and incubated with HRP-conjugated secondary antibodies (goat anti-mouse (H+L), 1:2,000, Invitrogen, cat. no. G21040; goat anti-rabbit, 1:3,000, Sigma-Aldrich, cat. no. 12-348) for 1 hour at room temperature. Following additional washes in TBS-T, immunoreactive bands were detected using enhanced chemiluminescence (Thermo Fisher Scientific, cat. no. PI3457) and visualized using a LI-COR Odyssey XF imaging system and the images were saved as TIFF files. Images of Western blot and Ponceau S staining data not shown in the main results figures are included in supplementary materials **(Fig. S3)**. Densitometric analyses were performed using ImageJ2 (version 2.9.0/1.53t). Western blot images were converted to 8- or 16-bit grayscale. Regions surrounding bands at the correct molecular weight for GCPII and GAPDH were manually selected and analyzed as the signal. Bands at other molecular weights were not included in the analyses. Lanes were selected sequentially, and intensity profiles were generated using the Gel Analyzer tool. Areas under the curve (AUCs) were measured using the wand tool following baseline normalization and exported for analysis. Band intensities were first normalized to denervation-duration-matched controls to obtain relative expression values and data were plotted using GraphPad Prism.

### Enzymatic Activity Assay

GCPII activity measurements were carried out based on modifications of previously published protocols (*76*). Muscle tissue was stored at -80 °C until GCPII enzyme activity measurements were performed. Samples were homogenized in ice-cold Tris buffer (40 mM, pH 7.5) containing protease inhibitors (Roche, Complete Protease Inhibitor Cocktail, Ref# 04693159001) by mechanical disruption on ice using a handheld homogenizer (Biomasher II) for 10 cycles of 10 seconds with 10 seconds between cycles) and further sonicated using Kontes’ Micro Ultrasonic Cell Disrupter (three pulses of 30 second duration, 30 second between pulses, and on ice). The resulting homogenates were spun down (16,000× g for two minutes at 4 °C), and the supernatants were collected for both GCPII activity and total protein analysis. The GCPII reaction was initiated upon the addition of cobalt chloride (1 mM) and 3H-NAAG (50.0 mCi/µmol). The reactions were carried out in 50 µL reaction volumes in 96-well microplates for 180 minutes at 37 °C. Reactions were terminated with ice-cold sodium phosphate buffer (100 mM, pH 7) with 1 mM EDTA. 90 µL aliquots from each terminated reaction were then transferred to 96-well spin columns containing ion-exchange resin and the plate centrifuged at 1000 rpm for eight minutes using a Beckman GS-6R centrifuge. NAA-[3H]-G was bound to the resin and [3H]-G eluted in the flow through. To ensure complete elution of [3H]-G, columns were washed twice with formic acid (1 M, 90 µL). The flow through and the washes were collected and 200 µL aliquots transferred to a solid scintillator-coated 96-well plate (Packard) and dried to completion. The radioactivity corresponding to [3H]-G was determined with a scintillation counter (Topcount NXT, Packard, counting efficiency 80%). Each sample was run in duplicate. Finally, the total protein was quantified per the manufacturer’s instructions using the Bio-Rad DC Protein Assay kit (Cat# 5000111), and GCPII activity data were calculated as femtomoles of NAAG hydrolysis per mg protein per hour (fmol/mg/h).

### Statistical Analyses

For biodistribution studies, tissue uptake was calculated in percent injected dose per gram (%ID/g), and groupwise comparisons within each timepoint were performed using Kruskal-Wallis test. For small animal PET/MR data, the ratio of operative leg SUVmean divided by non-operative leg SUVmean was compared between groups and across timepoints using repeated-measures ANOVA, in which group (i.e., unilateral sciatic nerve transection with repair versus transection without repair) was the between-subjects factor and timepoint (i.e., four weeks versus 16 weeks post-injury) was the within-subjects factor. Comparisons of pooled estimates of protein expression on Western blot and enzymatic activity between denervated samples and controls were performed using the Wilcoxon matched-pairs signed-rank test and the Mann-Whitney test, respectively. All statistical analyses were performed using GraphPad PRISM version 10.3.1 and SAS software version 9.4 (SAS Institute), and p-values < 0.05 were considered statistically significant.

## Data Availability

All data produced in the present study are available upon reasonable request to the authors

## Notes

### Competing Interest Statement

The authors have declared no competing interest.

### Funding Statement

This study was funded by grant support from the Department of Defense (DoD) and the American Society for Surgery of the Hand (ASSH).

### Author Declarations

The Institutional Review Board (IRB) of Johns Hopkins University gave ethical approval for this work (IRB00422704).

