## Supplemental Figures and Tables for "Glutamate Carboxypeptidase II (GCPII)-Targeted PET to Identify Muscle Denervation in Peripheral Nervous System Injuries"

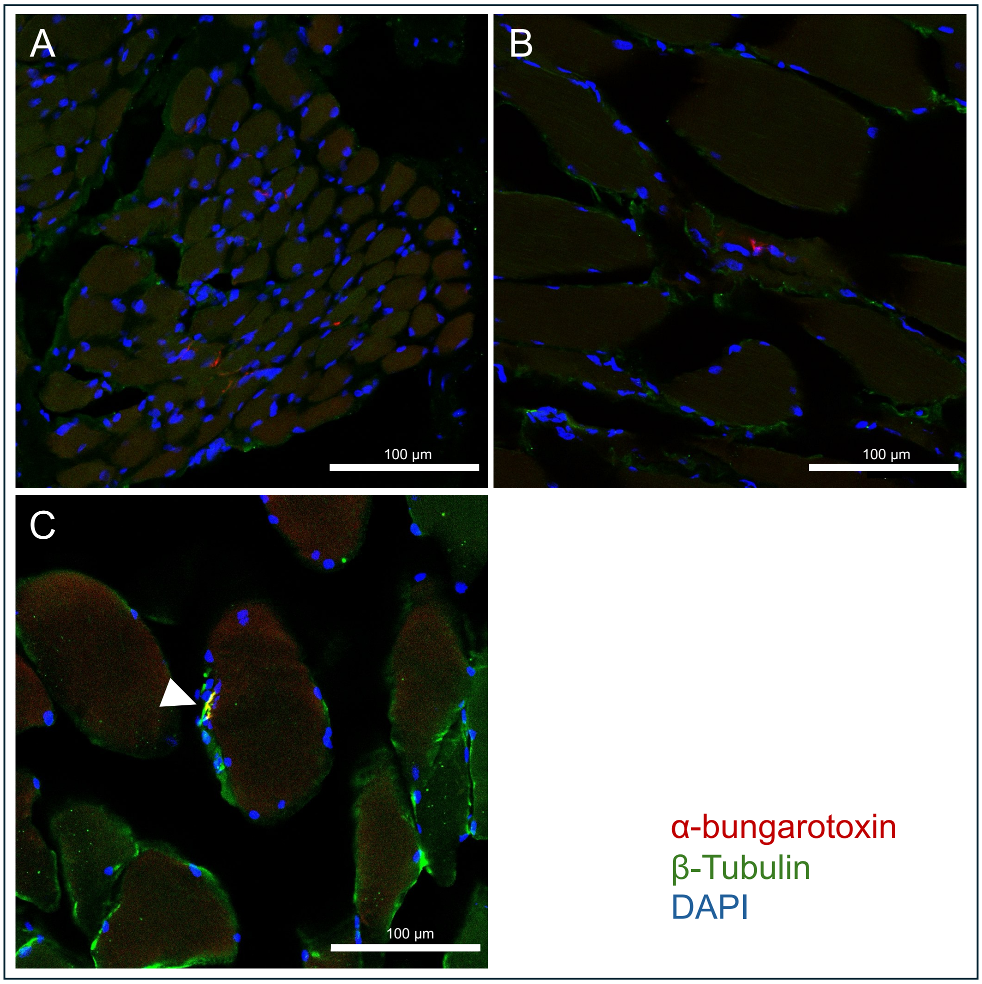


**Figure S1: Immunofluorescence demonstrating muscle denervation 16 weeks after porcine median nerve injury**. Composite images of α-bungarotoxin (red) targeting motor endplates, anti-βIII-tubulin (green) targeting axonal cytoskeletons, and DAPI (blue) targeting nuclei. Note lack of βIII-tubulin staining in the **(A)** left and **(B)** right flexor carpi radialis muscles at this timepoint, indicating denervation. In contrast, **(C)** the pectoralis major muscle (uninjured reference) demonstrates innervated neuromuscular junctions (arrowhead), indicated by co-localization between βIII-tubulin and α-bungarotoxin staining that appears yellow.

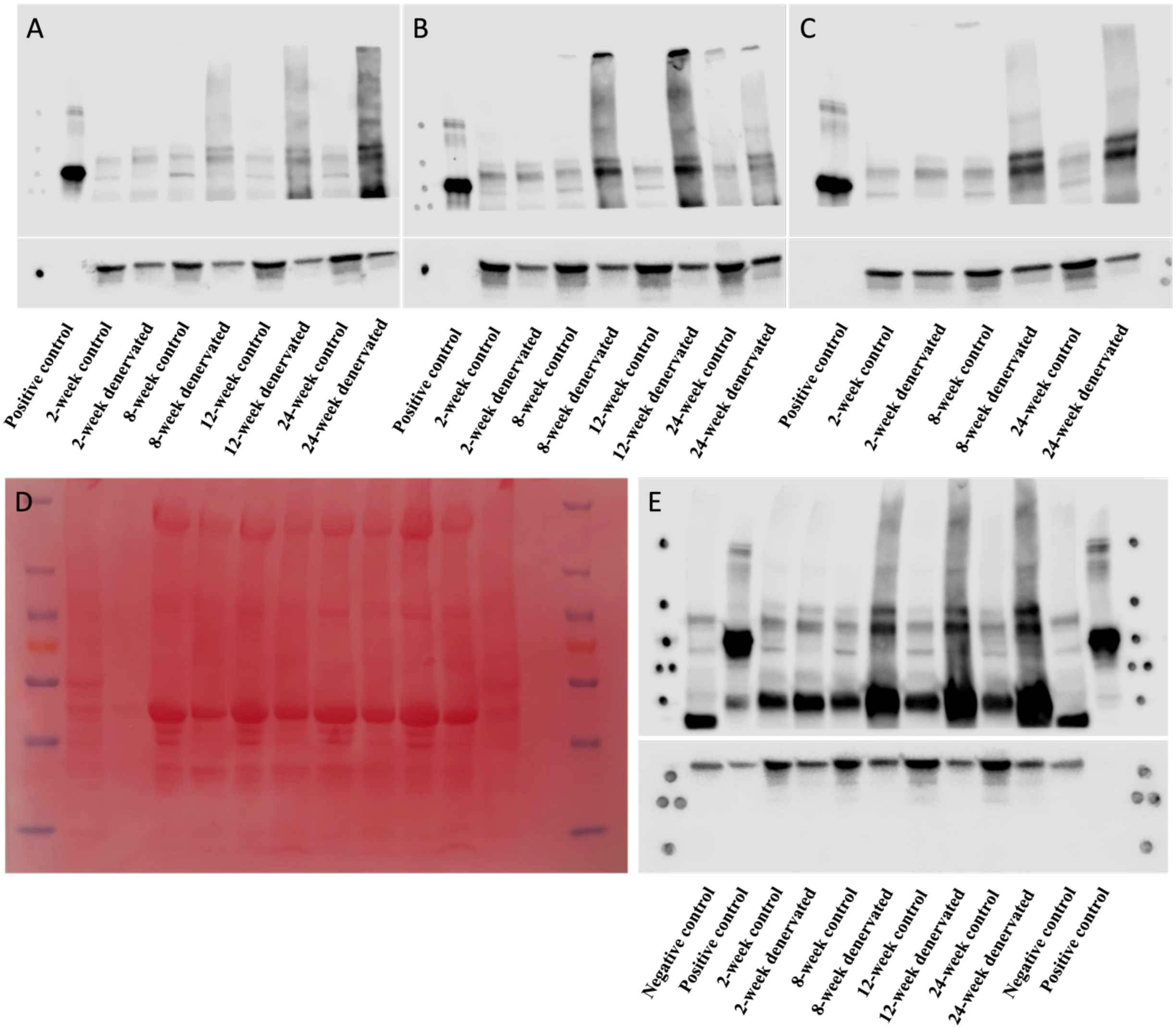


**Figure S2. (A, B, and C)** Individual Western blot gels showing GCPII expression in control and denervated rat lateral gastrocnemius muscle for each biological replicate (n=2–3 per group). Gels correspond to samples used for the densitometric analyses presented in Figure 3. **(D)** Ponceau S–stained membrane showing total protein loading. (E) Full, uncropped Western blot membrane corresponding to the representative GCPII blot in Figure 3.

**Table S1: Tissue [$^{68}$Ga]PSMA-11 uptake on porcine PET/CT**

| Tissue | Status | SUVmean |
|---|---|---|
| ***Deep Forearm Flexor Compartment*** | | |
| Left | Injured | 0.59 ± 0.12 |
| Right | Injured | 0.52 ± 0.12 |
| ***Superficial Forearm Flexor Compartment*** | | |
| Left | Injured | 0.45 ± 0.12 |
| Right | Injured | 0.39 ± 0.12 |
| Forearm Extensor Compartments | Unaffected | 0.30 ± 0.075 |
| Biceps Muscles | Unaffected | 0.32 ± 0.058 |
| Pectoralis Major Muscles | Unaffected | 0.28 ± 0.092 |
| Latissimus Muscles | Unaffected | 0.15 ± 0.058 |
| Liver | Reference organ | 0.52 ± 0.17 |

**Table S1. Tissue [$^{68}$Ga]PSMA-11 uptake on porcine PET/CT.** Both superficial and deep flexor compartments are affected by a median nerve injury. Note lower uptake in unaffected muscle groups. Activity within each muscle or compartment of muscles is expressed in SUVmean.

**Table S2: Tissue [$^{18}$F]DCFPyL uptake on human PET/CT**

| Tissue | Status | SUVmean |
|---|---|---|
| ***Mobile Wad Compartment*** | | |
| Left | Injured | 0.41 ± 0.045 |
| Right | Unaffected | 0.18 ± 0.022 |
| ***Forearm Extensor Compartment*** | | |
| Left | Injured | 0.41 ± 0.045 |
| Right | Unaffected | 0.18 ± 0.023 |
| ***Forearm Flexor Compartments*** | | |
| Left | Unaffected | 0.24 ± 0.025 |
| Right | Unaffected | 0.20 ± 0.028 |
| ***Triceps Muscle*** | | |
| Left | Unaffected | 0.22 ± 0.025 |
| Right | Unaffected | 0.18± 0.028 |
| ***Pectoralis Major Muscle*** | | |
| Left | Unaffected | 0.21 ± 0.026 |
| Right | Unaffected | 0.16 ± 0.026 |
| ***Thigh Anterior Compartment*** | | |
| Left | Unaffected | 0.16 ± 0.028 |
| Right | Unaffected | 0.16 ± 0.034 |
| ***Thigh Posterior Compartment*** | | |
| Left | Unaffected | 0.14 ± 0.040 |
| Right | Unaffected | 0.15 ± 0.034 |
| Liver | Reference organ | 1.88 ± 0.14 |
| Kidney | Reference organ | 11.82 ± 1.84 |

**Table S2: Tissue [$^{18}$F]DCFPyL uptake on human PET/CT**. The mobile wad compartment consists of the radial nerve-innervated brachioradialis, flexor carpi radials longus, and flexor carpi radials brevis muscles. The forearm extensor compartment consists of wrist and digital extensor muscles. The triceps muscle, while innervated by the radial nerve, receives innervation proximal to the site of injury. Note increased uptake in injured muscle groups when compared to unaffected muscles, including the left triceps muscle. Mean activity within each muscle or compartment of muscles is expressed in SUVmean.
